# Biochemical and Biophysical Characterization of Respiratory Secretions in Severe SARS-CoV-2 (COVID-19) Infections

**DOI:** 10.1101/2020.09.11.20191692

**Authors:** Michael J. Kratochvil, Gernot Kaber, Pamela C. Cai, Elizabeth B. Burgener, Graham L. Barlow, Mark R. Nicolls, Michael G. Ozawa, Donald P. Regula, Ana E. Pacheco-Navarro, Carlos E. Milla, Nadine Nagy, Samuel Yang, Stanford COVID-19 Biobank Study Group, Angela J. Rogers, Andrew J. Spakowitz, Sarah C. Heilshorn, Paul L. Bollyky

**Author notes:** Co-first authors (Michael J. Kratochvil, Gernot Kaber). Co-senior authors (Angela J. Rogers, Andrew J. Spakowitz, Sarah C. Heilshorn, Paul L. Bollyky). Correspondence to: Paul Bollyky, MD, PhD: Department of Medicine, Division of Infectious Diseases, Stanford University, 279 Campus Drive, Beckman Center, Stanford CA 94305, USA.

## Abstract

Thick, viscous respiratory secretions are a major pathogenic feature of COVID-19 disease, but the composition and physical properties of these secretions are poorly understood. We characterized the composition and rheological properties (i.e. resistance to flow) of respiratory secretions collected from intubated COVID-19 patients. We found the percent solids and protein content are all greatly elevated in COVID-19 compared to heathy control samples and closely resemble levels seen in cystic fibrosis (CF), a genetic disease known for thick, tenacious respiratory secretions. DNA and hyaluronan are major components of respiratory secretions in COVID-19 and are likewise abundant in cadaveric lung tissues from these patients. COVID-19 secretions exhibited heterogeneous rheological behaviors with thicker samples showing increased sensitivity to DNase and hyaluronidase treatment. These results highlight the dramatic biophysical properties of COVID-19 respiratory secretions and suggest that DNA and hyaluronan may be viable therapeutic targets in COVID-19 infection.

## Introduction

Severe infections of SARS-CoV-2, the virus responsible for the COVID-19 pandemic, can result in acute respiratory distress syndrome (ARDS)(1), a condition marked by viscous respiratory secretions and respiratory distress(2). The compositional and rheological properties of these respiratory secretions impair their mucociliary clearance, resulting in a build-up of fluids in the lungs during ARDS(3). This greatly inhibits oxygen exchange, often necessitating endotracheal intubation and mechanical ventilation(4). Treatments that target these respiratory secretions are desperately needed to improve clinical outcomes for COVID-19 patients as well as for other patients suffering from severe cases of ARDS. It is therefore important to understand the composition of these secretions to better guide treatment development efforts.

Levels of hyaluronan (HA), a linear glycosaminoglycan, are elevated in respiratory secretions in other forms of respiratory inflammation(5-9) including ARDS(10, 11). HA is produced at the cell surface by a variety of cell types(12) in response to viral DNA and other factors(13). HA is present in the body at molecular weights ranging from low kilodaltons to megadaltons(12, 14) and is known to have major effects on the viscoelasticity of respiratory secretions and other materials(15, 16). Additionally, HA plays important roles in innate immunity and antigenic responses in the lungs(17-20).

DNA levels are also elevated in some forms of respiratory inflammation(21, 22). This increase likely originates from dead cells, infiltrating neutrophils(23, 24), and potentially microbial contaminants(25, 26). Relatively small increases in DNA concentrations can dramatically change the rheological properties of a solution, a phenomenon that has been leveraged both naturally in the production of bacterial biofilms(27) and synthetically in the development of DNA-based hydrogels(28). In the context of lung infections, extracellular DNA has been suggested to increase viscosity of mucosal fluid and provide colonization opportunities for bacterial infections(21).

We hypothesized that DNA and HA are major contributors to the tenacious behavior of respiratory secretions from COVID-19 patients. To evaluate this premise, we sought to characterize the composition and rheological properties (e.g., viscosity and elasticity properties) of respiratory secretions from patients under mechanical ventilation due to severe COVID-19, given the importance of these parameters in other respiratory diseases(29, 30). As controls for these studies, we have included sputum samples from both healthy individuals and subjects with cystic fibrosis (CF), a genetic disease associated with notoriously thick lung secretions(31).

## Results

### Solids and proteins are increased in COVID-19 respiratory secretions

We collected respiratory secretions from ventilated COVID-19 patients, ranging from 5 to 70 years of age (Table 1) via suction catheter, with only a single sample from each individual included in the dataset. Respiratory secretion samples were collected from patients with CF via spontaneous expectoration and from healthy volunteers via sputum induction.

**Table 1.**
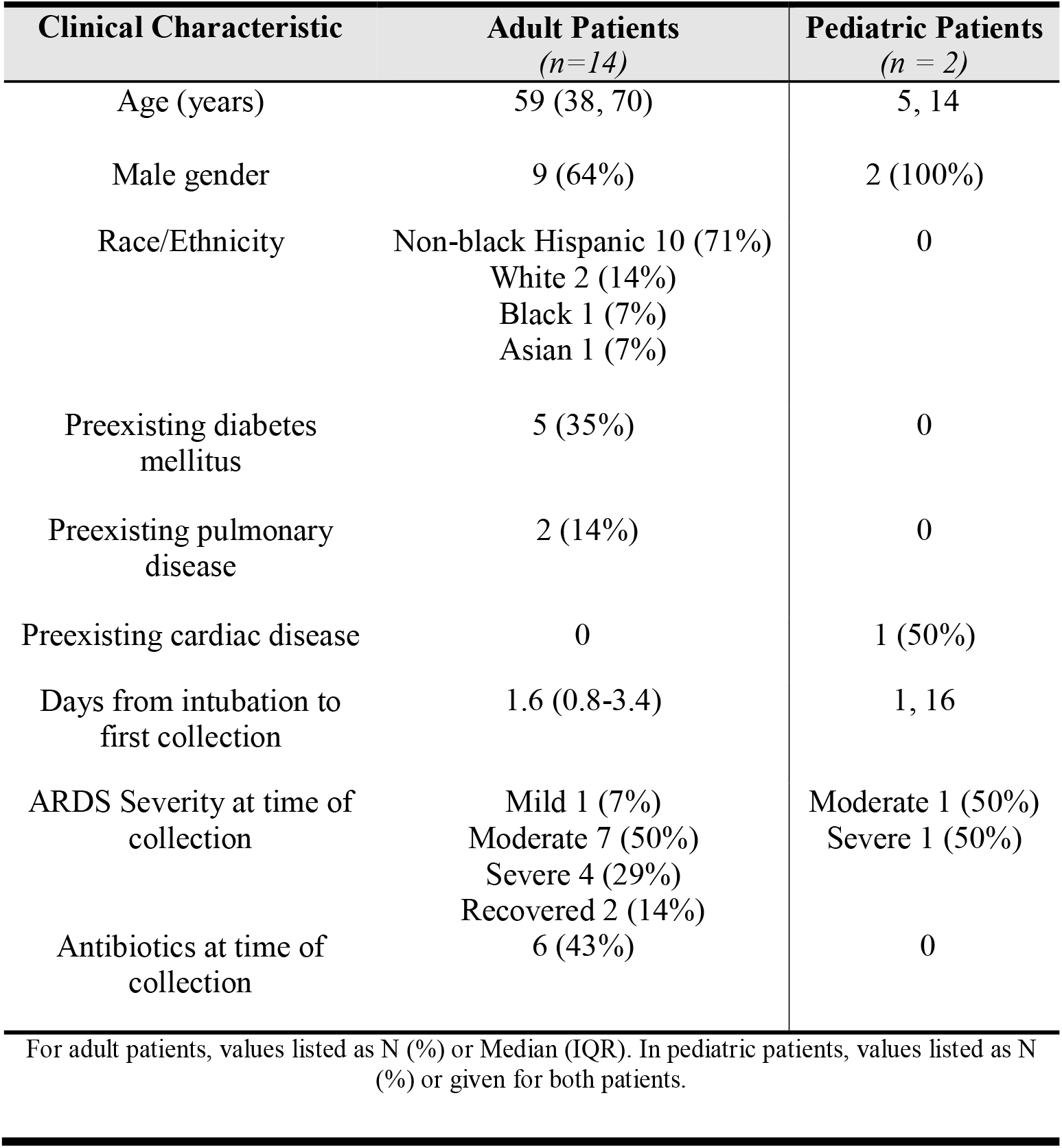
Clinical Characteristics of SARS-CoV2+ patients.

We observed that samples from healthy patients were typically clear and colorless, whereas samples from patients with COVID-19 were nearly always colored and opaque, similar to samples from patients with CF (Figure 1A), This suggested that the samples contain appreciable amounts of biopolymers and non-soluble debris.

**Figure 1.**
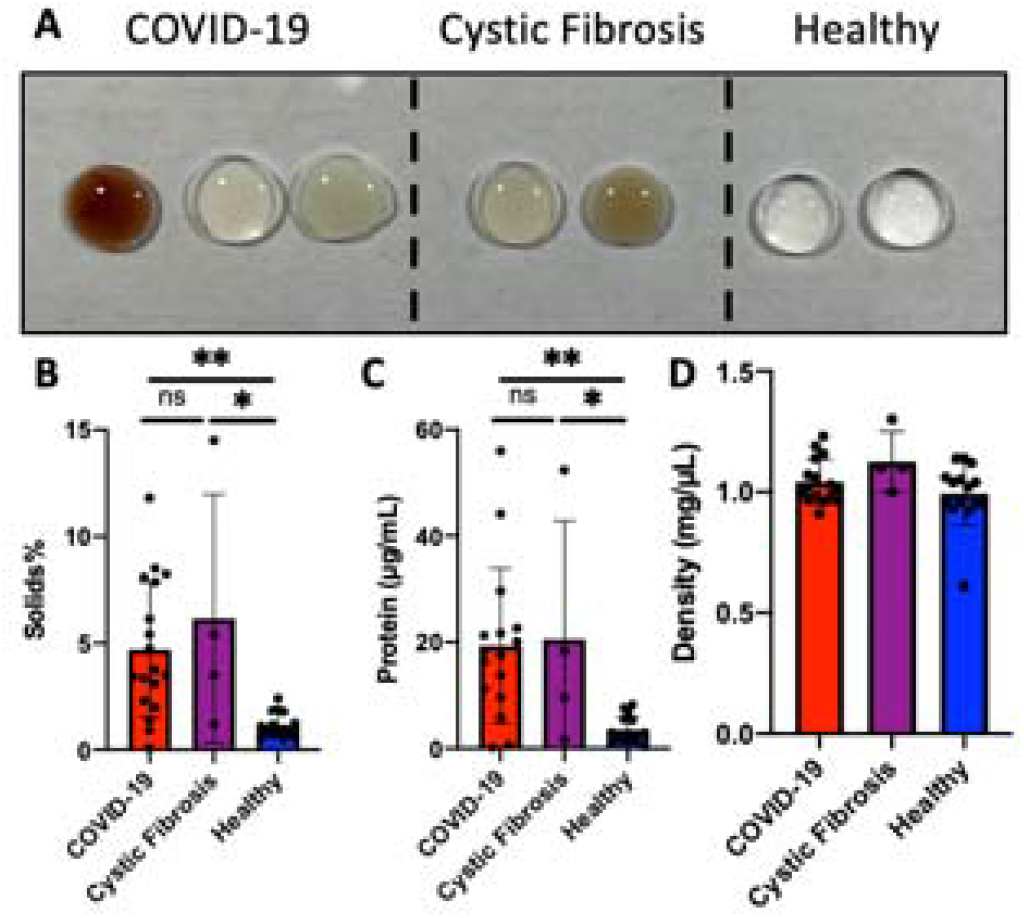
Respiratory secretions from patients with COVID-19 are high in solids and protein compared to healthy subjects. (A) Representative images of respiratory secretions collected from ventilated patients with COVID-19 are viscous and tenacious. Similar to CF samples, COVID-19 samples are often colored and opaque, whereas healthy samples are clear and colorless. (B) Quantification of solids found in COVID-19, CF, and healthy respiratory secretions. (C) Quantification of protein concentration in COVID-19, Cystic Fibrosis, and healthy respiratory secretions. (D) Density of respiratory secretions. One-way ANOVA with Tukey multiple comparisons tests. *p<0.05, **p<0.01.

The percent solids content of respiratory secretions, an index of hydration, impacts the difficulty with which respiratory secretions can be cleared and correlates with clinical outcomes in CF and other settings(32-34). We found that COVID-19 samples had significantly higher percent solids than healthy samples (Figure 1B). We further observed that protein concentrations in COVID-19 samples were nearly 5.5 times greater than those seen in healthy samples (Figure 1C, p = 0.003). COVID-19 and CF samples did not show statistically significant differences (p = 0.983). These data are consistent with infected and inflamed lungs being known to have protein deposits from increased mucin production(35), bacterial colonization(36), and infiltrating cells(24). However, there were no apparent changes in the density of the respiratory secretions (Figure 1D), suggesting that the contribution to density of the suspended and dissolved solids was not different enough to deviate from the density of healthy aspirate samples. Additionally, the pH of all samples was observed to be between 7-8.

Of note, large variances in solids and protein were observed in the COVID-19 patient samples (Figure 1B-C). Even though all the patient samples were collected from intubated patients with severe COVID-19 relatively early during mechanical ventilation, this variance may reflect the differences in the individual response to the infection and the disease progression at the time of collection.

### HA is increased in COVID-19 respiratory secretions and lung sections

We next evaluated HA content in the respiratory secretions. We observed a statistically significant, 10-fold increase in HA concentration in COVID-19 patient samples compared to samples from healthy subjects (Figure 2A, p = <0.0001). The average concentration of HA found in samples from COVID-19 subjects was comparable to that observed in samples from CF subjects (p = 0.333), a disease state associated with greatly increased sputum HA(37). Similar to our findings with percent solids and protein, we observed larger variance in the amounts of HA in COVID-19 and CF patient samples than compared to samples from healthy donors.

**Figure 2.**
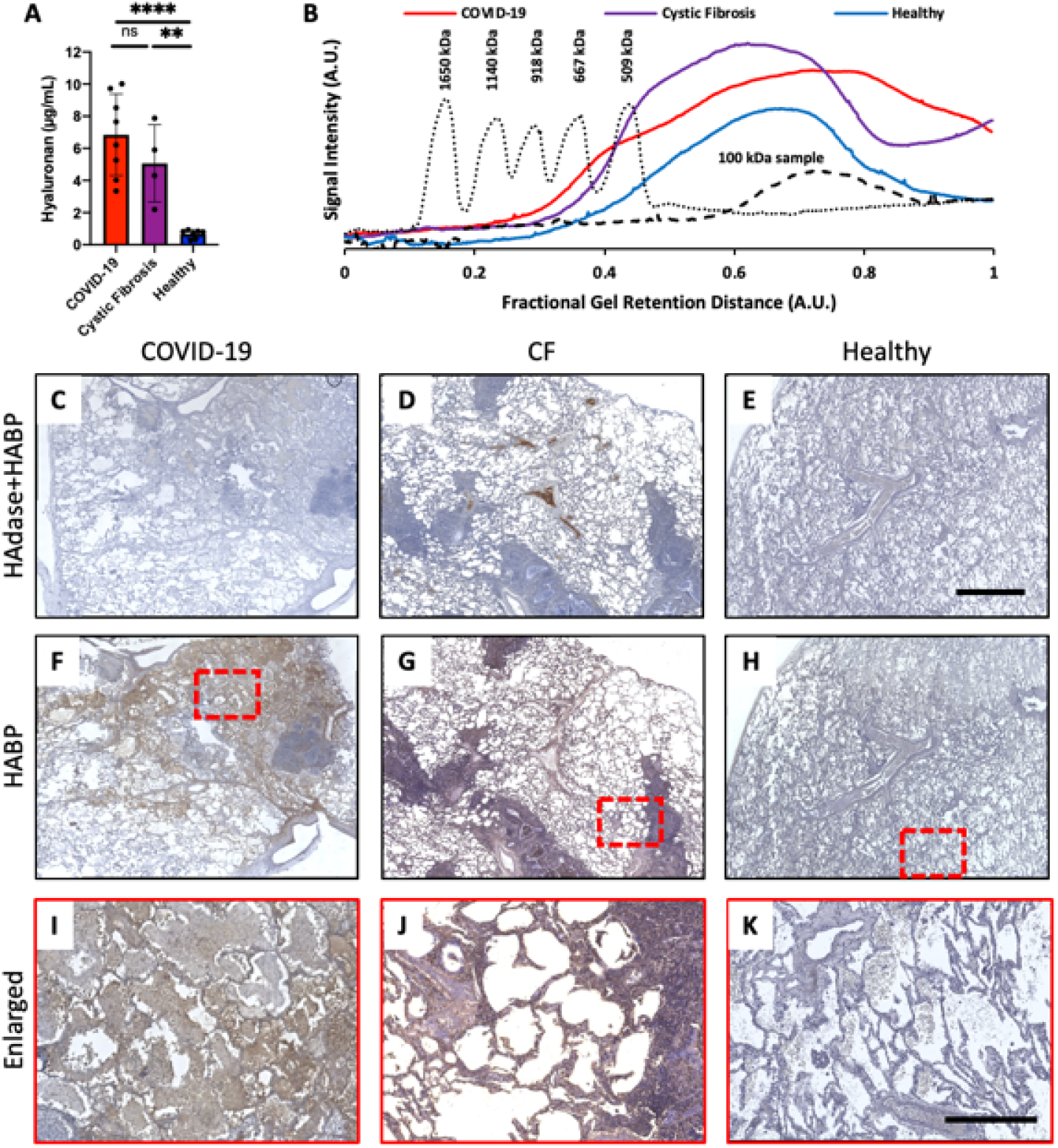
COVID-19 Patients have high levels of HA. (A) Quantification of HA in respiratory secretion samples. One-way ANOVA with Tukey multiple comparisons tests; **p<0.01, ****p<0.0001. (B) Representative chromatogram of HA molecular weight. Solid traces are the averages of COVID-19, CF, and healthy respiratory secretion samples. The dotted trace is the chromatogram of standard loaded with HA of known MWs, as indicated on graph. The dashed trace is representative of a commercially available 100 kDa MW HA. (C-H) Representative histological cadaveric lung sections from donors with COVID-19, donors with CF, and healthy donors, both with (C-E) and without (F-H) HAdase treatment. Nuclei are stained in blue, and HA binding proteins (HABP) are stained in brown. (I-K) Enlarged sections from panels F-H, respectively. Scale bars C-H 800 µm, I-K 400 µm.

Given that the molecular weight of HA is known to influence both the immunogenic as well as the rheological properties of the resulting solution(38-40), we measured the molecular weight of the HA found in the different samples (Figure 2B; Supplemental Figure 1). We found that while all samples of respiratory secretions had HA of molecular weight less than 500 kDa, HA size in samples from patients with COVID-19 donors skewed smaller than that seen in samples from donors with CF and healthy controls. Given that low molecular weight HA polymers promote inflammation in some systems(38, 39) this is consistent with the highly inflammatory nature of respiratory disease in COVID-19 infection.

### HA is increased in COVID-19 cadaveric lung sections

We next examined cadaveric lung sections from patients with COVID-19, patients with CF, and patients with healthy lungs (i.e., without a diagnosed pulmonary disease) for HA deposits by staining with HA binding protein (HABP). We observed very little HABP staining in sections treated with hyaluronidase (HAdase) (Figure 2C-E), suggesting very low non-specific background staining. However, we observed a substantial increase in HA staining in lung sections from both COVID-19 and CF donors compared to healthy samples when no prior HAdase treatment was used (Figure 2F-H). Higher magnification images demonstrated the accumulation of HA within alveolar spaces (Figure 2I-K). These data, together with the aforementioned respiratory secretion studies, indicated that the respiratory secretions of patients with severe COVID-19 have elevated levels of HA.

### DNA is increased in COVID-19 respiratory secretions

We next examined the double-stranded DNA (dsDNA) content in these respiratory secretion samples. We observed that the respiratory secretions collected from patients with COVID-19 had a statistically significant increase in dsDNA content compared to healthy subjects (Figure 3A, p = 0.032). The average dsDNA content in the COVID-19 samples was 14 times greater than that in the healthy samples, with eight of the observed samples having over ten times more dsDNA. CF sputum, by comparison, had roughly a comparable average dsDNA concentration to COVID-19 samples (p = 0.999). Sizing the dsDNA in the samples suggested that the dsDNA is very large (greater than 10 kb, i.e. >6,000 kDa) (Figure 3B; Supplemental Figure 2). The variance observed in samples from patients with COVID-19 and CF was again very large in comparison to that observed in healthy donor samples.

**Figure 3.**
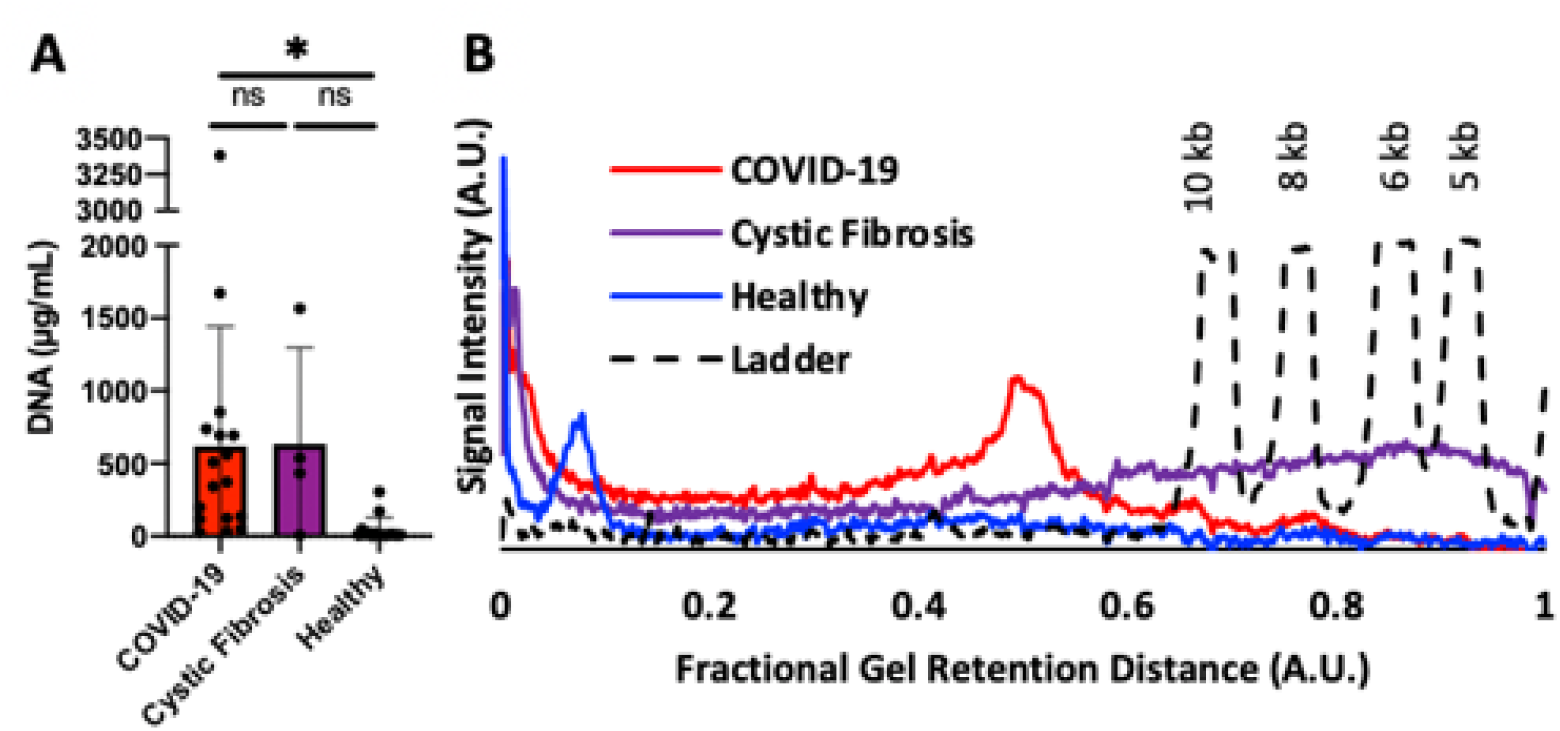
Increased levels of dsDNA in COVID-19 respiratory secretions. (A) Quantification of dsDNA in respiratory secretion samples. One-way ANOVA with Tukey multiple comparisons tests. *p<0.05. (B) Representative chromatogram of dsDNA molecular weight. Solid traces are the averages of COVID-19, CF, and healthy respiratory aspirate samples. The dashed line trace is the chromatogram of a DNA standard ladder with the dsDNA base pair-lengths labeled above the respective peaks.

### High modulus secretions are more susceptible to enzymatic treatment

We evaluated the rheological properties of COVID-19 respiratory secretions and the contribution of HA and dsDNA to the physical flow properties of the secretions. These flow properties are of crucial importance for the removal of the secretions from the lungs. Dynamic light scattering microrheology, a non-invasive rheology technique, was used to evaluate the rheological properties of the sample due to the small sample volume required and the ability of the technique to not alter the sample properties during measurement(41, 42). The samples were measured both before and following enzymatic treatment (microrheology protocol further described in Methods and Supplemental Figures 3-4). Specifically, we examined the impact of enzymatic treatment with HAdase (to degrade HA) or deoxyribonuclease(43) (DNase; to degrade dsDNA) on the flow properties of respiratory secretions, as we hypothesized that enzymatic degradation of these biopolymers would lower the modulus (i.e. the resistance to flow). As a non-enzymatic treatment control, samples were diluted with an equivalent volume of saline. When compared to dilution, the average modulus relative to pre-treatment was lower following enzymatic treatment with either HAdase or DNase, although the comparison was not statistically significant (Supplemental Figure 4). However, we suspected the lack of significance was largely impacted by the wide variance of pre-treatment moduli of the secretions.

To account for this large variance in pre-treatment samples, we then evaluated the impact of enzymatic treatment as a function of the measured pre-treatment modulus of the respiratory secretions (Figure 4). As expected, samples that had a higher pre-treatment modulus (i.e. thicker samples that were more resistant to flow) had a larger response to enzymatic treatment by either DNase or HAdase compared to a control saline dilution. We found a statistically significant linear relationship between the pre-treatment modulus of the secretions and the difference between the change of modulus with dilution and change of modulus with enzymatic treatment (ΔG_Saline_ – ΔG_Enzyme_). If the enzyme had no effect compared to the dilution control, then ΔG_Saline_ – ΔG_Enzyme_ = 0; if the enzyme treatment decreased the modulus of the sample, then ΔG_Saline_ – ΔG_Enzyme_ < 0. The Bayesian Information Criterion for comparing this linear model against a fit for random noise was much greater than 10 (BIC = 35.75 for DNase, and BIC = 36.92 for HAdase), indicating very strong statistical significance for this linear relationship. These data are consistent with the hypothesis that thicker COVID respiratory secretions are sensitive to enzymatic treatments that degrade DNA and HA, resulting in a lower modulus (i.e. less resistance to flow). By comparison, healthy control samples had low pre-treatment moduli and were not dramatically impacted by enzymatic treatments.

**Figure 4.**
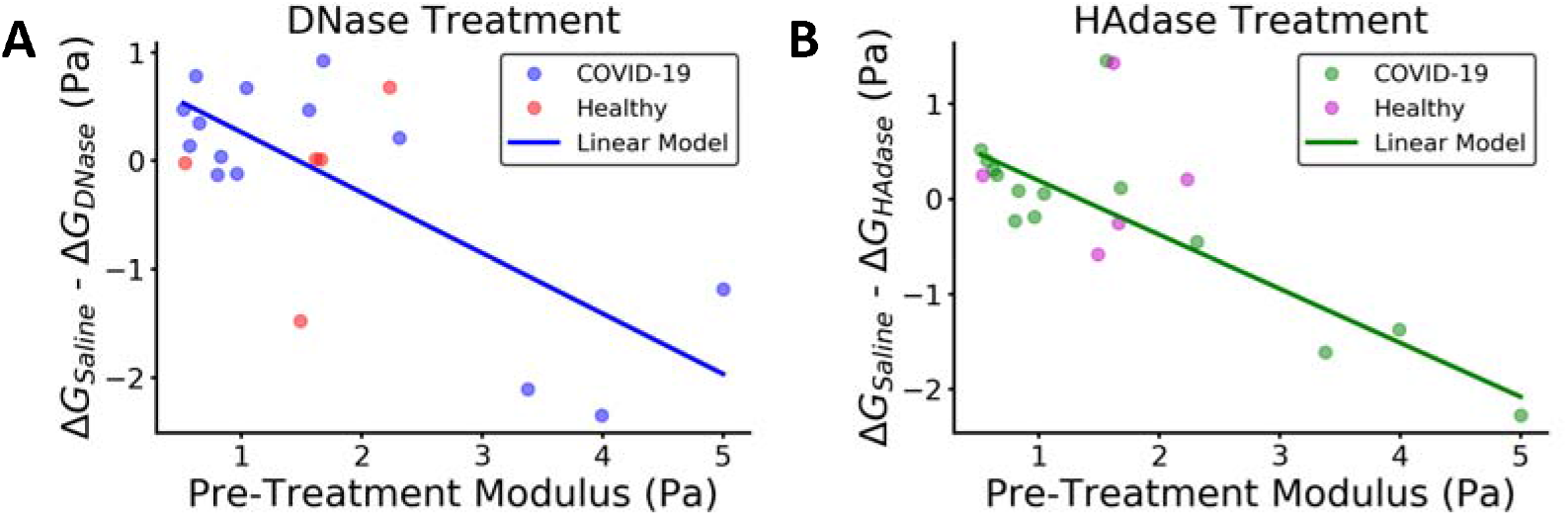
Enzymatic treatments impact the rheology of COVID-19 respiratory secretions in proportion to their pre-treatment moduli. (A) The difference in modulus of COVID-19 lung secretions upon control saline dilution and enzymatic DNase treatment (ΔG_Saline_-ΔG_DNase_) versus the initial, pre-treatment modulus (blue). Healthy controls shown for comparison (red). (B) The difference in modulus of COVID-19 lung secretions upon control saline dilution and enzymatic HAdase treatment (ΔG_Saline_-ΔG_HAdase_) versus the initial, pre-treatment modulus (green). Healthy controls shown for comparison (magenta).

## Discussion

Here, COVID-19 respiratory secretions were observed to be similar to the notoriously thick and tenacious sputum produced by patients with CF. COVID-19 respiratory secretions have significantly elevated levels of solids, with HA and DNA contributing to the elevated viscosity. Low-molecular weight HA is greatly increased in the respiratory secretion samples from intubated patients with COVID-19. Consistent with this, HA is abundant in histologic sections from cadaveric lung tissue obtained from an individual with COVID-19-associated ARDS. Together these data support the hypothesis that low-molecular weight HA is elevated in the respiratory secretions of patients with COVID-19-associated ARDS and may be a contributing factor in the inflammatory state in the lungs.

Due to their potential impact on the flow properties of the respiratory secretions, we further evaluated the rheological properties of the respiratory secretions in response to enzymatic degradation of HA and DNA. Similar to the variance observed in the solids and protein content of COVID-19 respiratory secretions, the rheological values of these secretions also exhibited large variance. The typical composition of respiratory secretions consists of dilute mucins(44), which are long polymers that can form a network by entangling with each other within the aspirate fluid. Similar to mucin, other biopolymers, such as DNA and HA, can form entanglements with itself and with mucin, forming more entanglements with increasing polymer concentration and contributing to a greater modulus (i.e., a greater resistance to flow). Respiratory secretions with higher modulus are expected to be more challenging to clear from the airway and hence hinder oxygen exchange in the lungs(4, 26, 45). Thus, thinning of the fluid to improve lung clearance is a common goal across a range of diseases with respiratory inflammation(46-49). As we observed in our study, treatment of respiratory secretions with an enzyme to digest the biopolymers (and hence decrease the polymer entanglements) will decrease the flow resistance of thick samples with an initial high modulus. The impact of the DNase is established clinically, as it has been used in treating CF lung disease(50, 51); however, the use of HAdase for improving the flow properties of respiratory secretions is a relatively new approach and requires further investigation. More research is needed to identify ideal treatment conditions such as dosages and dosage regimens. Further, targeting the production of the HA during infection may be more successful than relying on a post-production degradation approach. Treatment with a pharmaceutical HA-inhibitor such as 4-methylumbiliferone(52) may be a viable approach to limit the deposition of HA during infection.

These studies have several limitations. Most notable is the small numbers of cases and samples of secretions involved. These findings need to be confirmed in larger, multi-center studies involving individuals with diverse backgrounds and case presentations. The underlying mechanisms that lead to increased HA would also benefit from further research to identify the causative cell types and signalling pathways. In addition, data in SARS-CoV-2 animal models would enable improved understanding of the contribution of HA to pathogenesis in this disease. Finally, to safely acquire the rheology data, the COVID-19 samples were heat treated to render the samples non-infectious. In control CF samples, this same heat treatment was found to decrease the modulus (Supplemental Figure 4A), presumably due to the denaturation of biopolymers in the sample. Since we observed that higher modulus samples had larger responses to enzymatic treatment, the true effect of enzymatic treatment on COVID-19 lung secretion is likely larger than that reported here using heat-treated samples. Based on the promising data presented here, future studies should further evaluate a range of enzymatic treatment dosages and durations to assess the rheological effects on non-heat-treated COVID-19 lung secretions.

In summary, these data indicate elevated dsDNA and HA levels in COVID-19 respiratory secretions. These studies may have important implications for the development of much needed therapeutics for patients with COVID-19. Developing treatments that render the respiratory secretions of lungs less viscous, and thus easier to clear via natural mucociliary clearance, could be pivotal to improving clinical outcomes in severe COVID-19 and ARDS.

## Methods

### Histologic staining of lung tissues for HA

Human lung tissue was obtained from a de-identified autopsy specimen provided through the Stanford Pathology Department in the form of formalin-fixed, paraffin-embedded histologic specimen. Histological staining for HA was performed as described previously(53). In brief, 5-µm thick sections were cut on a Leica RM 2255 Microtome (Leica Microsystems Inc.). For HA affinity histochemistry (AFC) the Bond Intense R Detection kit, a streptavidin-horse radish peroxidase (HRP) system, (Leica Microsystems, Inc.) was used with 4 µg/mL biotinylated-HABP in 0.1 % bovine serum albumin (BSA) in phosphate buffered saline (PBS) as the primary. All images were collected using the BZ-X710 inverted fluorescence microscope (Keyence, Osaka, Japan) at 20X magnification. Montages were generated using the Keyence BZX Analyzer software’s stitching function.

### Collection of human respiratory secretions

We collected respiratory secretions from patients enrolled in the Stanford University sputum biobank study from March–November 2020. COVID-19 samples were discarded respiratory secretions obtained during the course of routine clinical care. All COVID-19 samples were collected from ventilated patients who were diagnosed with ARDS. Eligibility criteria included admission to Stanford Hospital with a positive SARS-CoV-2 nasopharyngeal swab by RT-PCR. Patients admitted to the ICU were included. Patients were phenotyped for ARDS using the Berlin criteria (acute onset of hypoxemic respiratory failure with a PaO2/FIO2 ratio (i.e., the ratio of the partial pressure of arterial oxygen to the percentage of inspired oxygen) of <300 on at least 5 cm of positive end-expiratory pressure, bilateral infiltrates on chest X-ray). For controls, sputum was collected from asymptomatic adult donors. Healthy control subjects were asymptomatic, aged 24–50 years. Sputum samples from CF patients were collected during routine care. All samples were frozen at -80°C immediately after collection. Samples were thawed slowly on ice then heat treated at 65°C for 30 minutes to render the virus inactive and the sample noninfectious prior to further analyses. These studies were approved under APB protocol #2379.

### Compositional characterization of respiratory secretions

The solids content of the human respiratory secretions was determined by taking the ratio of the freeze-dried mass and wet mass of the samples following at least 2 days of lyophilization. Protein concentrations were determined using the Pierce Bicinchoninic Acid (BCA) Protein Assay (Thermo Scientific) following manufacturer’s instructions. HA concentration was determined using a modified HA Enzyme-Linked Immunosorbent Assay (ELISA) as previously described(54). DNA concentrations were determined using Quant-iT dsDNA Broad-Range Assay Kit (Molecular Probes-Life Technologies) following manufacturer’s instructions. The pH of the samples was measured using pH-indicator strips (Supelco).

### Gel electrophoresis to characterize HA molecular weight

Respiratory secretion samples were treated with 250 U benzonase for 30 min at 37°C for nucleic acid digestion, followed by an incubation with 1 mg/ml proteinase K for 4 hrs at 65°C for further digestion. Proteinase K was heat inactivated by incubating the samples at 100°C for 5 min. Insoluble material was removed by centrifugation at 17,000 g for 10 min before further processing. Samples were precipitated with ethanol overnight at -20°C by adding 4 volumes of pre-chilled 200-proof ethanol to each sample. The following day, the samples were centrifuged at 17,000 g for 10 min. The supernatant was discarded, and the pellet was washed by adding 4 volumes of pre-chilled 75% ethanol. Samples were centrifuged at 17,000 g for 10 min and the resulting pellet air dried at room temperature for 20 minutes. Each sample was resuspended in 100 μl of 100 mM ammonium acetate in water, lyophilized and resuspended in 10 μl of 10 M formamide. Samples were separated on a 1% Tris-acetate EDTA (TAE) agarose gel run at 100 V, then stained with Stains-All (1.25 mg/200 mL in 30% ethanol) (Sigma). The gel was imaged on a BioRad GS-800 Calibrated Densitometer. Twice the volume of healthy control samples was loaded in each gel lane compared to CF and COVID-19 samples.

### Gel electrophoresis to characterize DNA molecular weight

Respiratory secretion samples were mixed with loading solution and separated on a 1% agarose gel with 0.5 µg/mL ethidium bromide at 120 V. Samples were separated on the same gel with a 2-Log DNA Ladder (New England Biolabs). Gels were imaged on a BioRad ChemiDoc MP imaging system.

### Microrheology measurements

Dynamic light scattering (DLS) microrheology data was collected as previously described^41,42^ with minor modifications as described below. Due to the presence of naturally occurring particulates within all samples, no additional beads were required to induce light scattering. Light scattering was collected from a Malvern Nano Zetasizer Nano ZS with a 633 nm laser operated in 173º backscatter mode. The raw intensity autocorrelation function of a respiratory secretion sample was measured at a specified measurement position for 30 minutes at 37ºC. Following initial microrheology measurements, the same respiratory secretion sample was then treated with either 1) benzonase nuclease (250 U/mL) for 1 hour at 37°C, 2) hyaluronidase (50 mg/mL, Sigma Aldrich) for 2 hour at 37ºC, or 3) 1x phosphate buffered saline for 1 hour at 37ºC as a dilution control. After the allotted reaction time, the DLS measured the raw intensity autocorrelation function of the sample at the same settings as before. To safely determine the effect of heat on the rheological behavior of respiratory secretions, we measured the intensity autocorrelation function of CF sputum, which is similar to COVID-19 respiratory secretions in both composition and rheological behavior, before and after the same heat treatment that all COVID-19 respiratory secretion samples were subjected to prior to handling. The heat treatment significantly decreased the resistance to flow (i.e. the elastic modulus) of the CF sputum (Supplemental Figure 4A).

### Microrheology data analysis

The intensity autocorrelation data acquired above was analyzed using the custom analysis package found at dlsur.readthedocs.io. The size of the particulates was assumed to be 500 nm in diameter for all samples. While this assumption affects the absolute value of the modulus derived from the scattering autocorrelation function, it has the same proportional effect across all samples. Thus, the trends observed in the microrheology data, along with the conclusions drawn from those trends, are unaffected by this assumption. All rheological measurements in this study obtained the complex modulus over a wide range of frequencies (from about 10^1^ to 10^6^ Hz), but only the modulus value at one frequency was used when comparing the modulus across samples and conditions. This is a common approach when comparing rheological results of lung secretions(21). To determine this frequency, the complex moduli of the pre-and post-treatment were compared. In the spectrum with the higher complex modulus (typically the pre-treatment), a single frequency was determined by selecting either a) the middle of the “plateau” region of the elastic modulus (Supplemental Figure 3A) or, in the case of no plateau region, b) the lowest frequency for which there is data (Supplemental Figure 3B). Some samples had limiting frequency ranges due to the fast decay of the measured autocorrelation function, which often corresponds to solutions with less resistance to flow. For a single sample, the same frequency was chosen for the pre-treatment modulus and post-treatment modulus. The change in modulus with dilution, ΔG_Saline_, was determined by subtracting the modulus of the sample after dilution to before dilution. The change in modulus with enzyme (DNase or hyaluronidase) treatment, ΔG_enzyme_, was determined by subtracting the modulus of the sample after enzyme addition from the modulus before enzyme addition. The measured moduli pre-treatment, G_pre_, and post-treatment, G_post_, for all individuals are shown in Supplemental Figure 4B.

### Statistics

Data are expressed as mean +/- SEM of n independent measurements. Significance of the difference between the means of two or three groups of data was evaluated using a one-way ANOVA followed by Tukey’s post hoc test. A p value less than <0.05 was considered statistically significant. The linear model fit used to compare enzyme effects on rheological properties of different samples was evaluated using the Bayesian Information Criterion. The Bayesian Information Criterion for a linear fit and a random model fit were compared, and a Bayesian Information Criterion above 10 was considered to be very strong evidence against the random model.

## Data Availability

All primary data is available upon request.

## Data Availability

All data generated and analyzed during the current study are available from the corresponding author upon reasonable request.

## Study Approval

All secretion samples were obtained under the auspices of research protocols approved the Stanford Institutional Review Board (IRB) (Stanford IRB approval #28205, #53685, #55650, #37232, and #43805). Samples were collected after written informed consent from patients or their surrogates prior to inclusion in the study.

## Author contributions

G.K., P.L.B., A.J.S, and S.C.H. conceived the study. E.B.B, M.R.N., M.G.O., D.P.R., A.E. P.-N., Stanford COVID Biobank, C.E.M., and A.J.R. identified, enrolled, and consented eligible patients and patient samples. M.J.K., G.K., and P.L.B. processed patient samples. M.J.K., G.K., P.C.C., G.L.B., and N.N. performed experiments and analyses. A.J.S. performed data analysis. M.J.K., G.K., P.C.C., E.B.B., M.R.N, N.N., A.J.R., A.J.S., S.C.H., and P.L.B. interpreted data and wrote the manuscript with input from all authors. Authorship order for co-first authors was determined via mutual agreement between M.J.K. and G.K.

## Acknowledgements

We are grateful to all participants in this study. Our thanks go to A. Wardle for his reading of the manuscript and his helpful comments. We would also like to thank G. Nolan for providing access to the Keyence microscope and analysis software. This research program is supported by a grant from the Stanford Innovative Medicines Accelerator program and the COVID-19 Response program from Stanford ChEM-H (Chemistry, Engineering & Medicine for Human Health). The Stanford COVID-19 Biobank Study Group and A.J.R. are funded by NIH/ NHLBI K23 HL125663. A.J.R. was supported by NIH grant T32 AI007502-23. See Supplemental Acknowledgments for Stanford COVID-19 Biobank Study Group details.

## Materials & Correspondence

Please address correspondence and materials requests to Paul Bollyky, MD, PhD: Department of Medicine, Division of Infectious Diseases, Stanford University, 279 Campus Drive, Beckman Center, Stanford CA 94305, USA.

## Notes

**Conflict of Interest:** The authors have declared no conflict of interest exists.

### Competing Interest Statement

The authors have filed a patent on the use of 4-methylumbelliferone, an inhibitor of hyaluronan synthesis in acute respiratory distress syndrome and COVID-19 infection.

### Funding Statement

This research program is supported by a grant from the Stanford Innovative Medicines Accelerator program. The Stanford ICU Biobank and A.J.R. are funded by NIH/ NHLBI K23 HL125663. A.R. was supported by NIH grant T32 AI007502-23.

### Author Declarations

Stanford University IRB

