## Supplemental files for "Biochemical and Biophysical Characterization of Respiratory Secretions in Severe SARS-CoV-2 (COVID-19) Infections"

Michael J. Kratochvil^1,2#^, Gernot Kaber^1#^, Pamela C. Cai^3^, Elizabeth B. Burgener^4^, Graham L. Barlow^1^, Mark R. Nicolls^5^, Michael G. Ozawa^6^, Donald P. Regula^6^, Ana E. Pacheco-Navarro^5^, Carlos E. Milla^4^, Nadine Nagy^1^, Samuel Yang^7^, Stanford COVID-19 Biobank Study Group, Angela J. Rogers^5*^, Andrew J. Spakowitz^3*^, Sarah C. Heilshorn^2*^ and Paul L. Bollyky^1*^

**Table of Contents**

Supplemental Figure 1. HA molecular weight sizing gel

Supplemental Figure 2. DNA molecular weight sizing gel

Supplemental Figure 3. Pre-treatment and post-treatment modulus identification

Supplemental Figure 4. Microrheology data analysis

Supplemental Acknowledgments

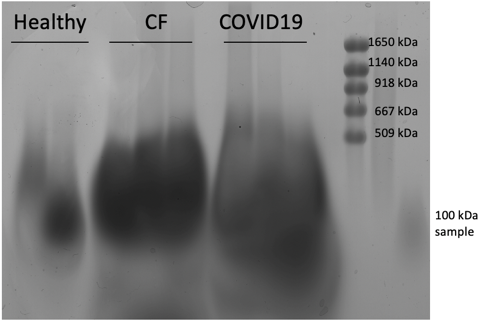

**Supplemental Figure 1.** Representative HA molecular weight sizing gel from Healthy, CF, and COVID-19 respiratory secretion samples.

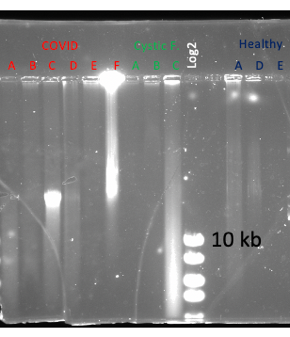

**Supplemental Figure 2.** Representative DNA molecular weight sizing gel from Healthy, CF, and COVID-19 respiratory secretion samples.

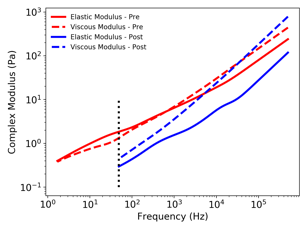

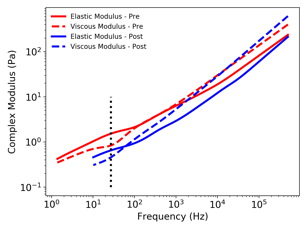

**A**

**B**

**Supplemental Figure 3.** Representative rheological spectra for pre-treatment and post-treatment samples, where the dotted line represents the frequency chosen at which the single modulus value was used for comparison when (A) a "plateau" region of the elastic modulus exists or, (B) in the case of no plateau region and hence the lowest frequency is selected.

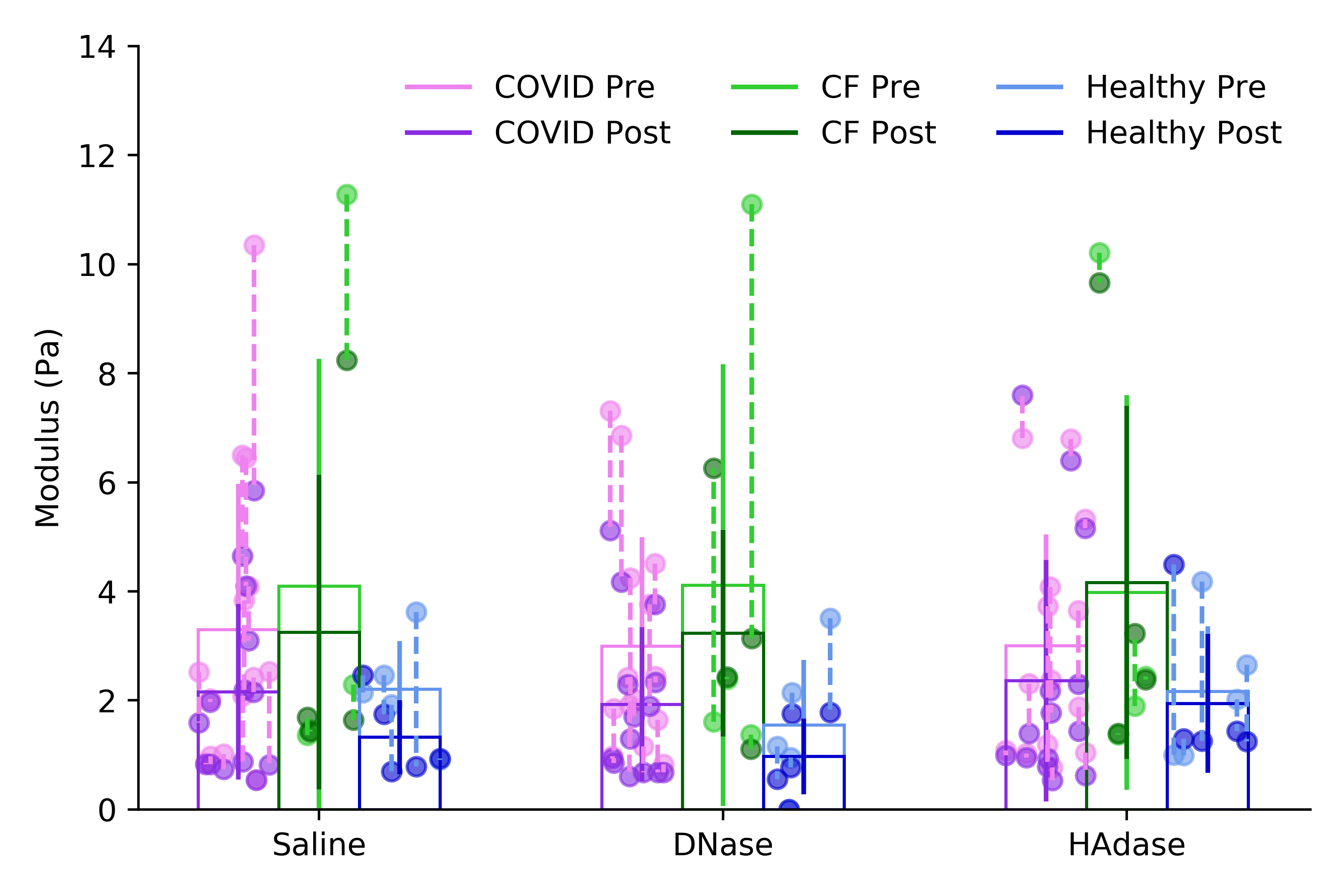

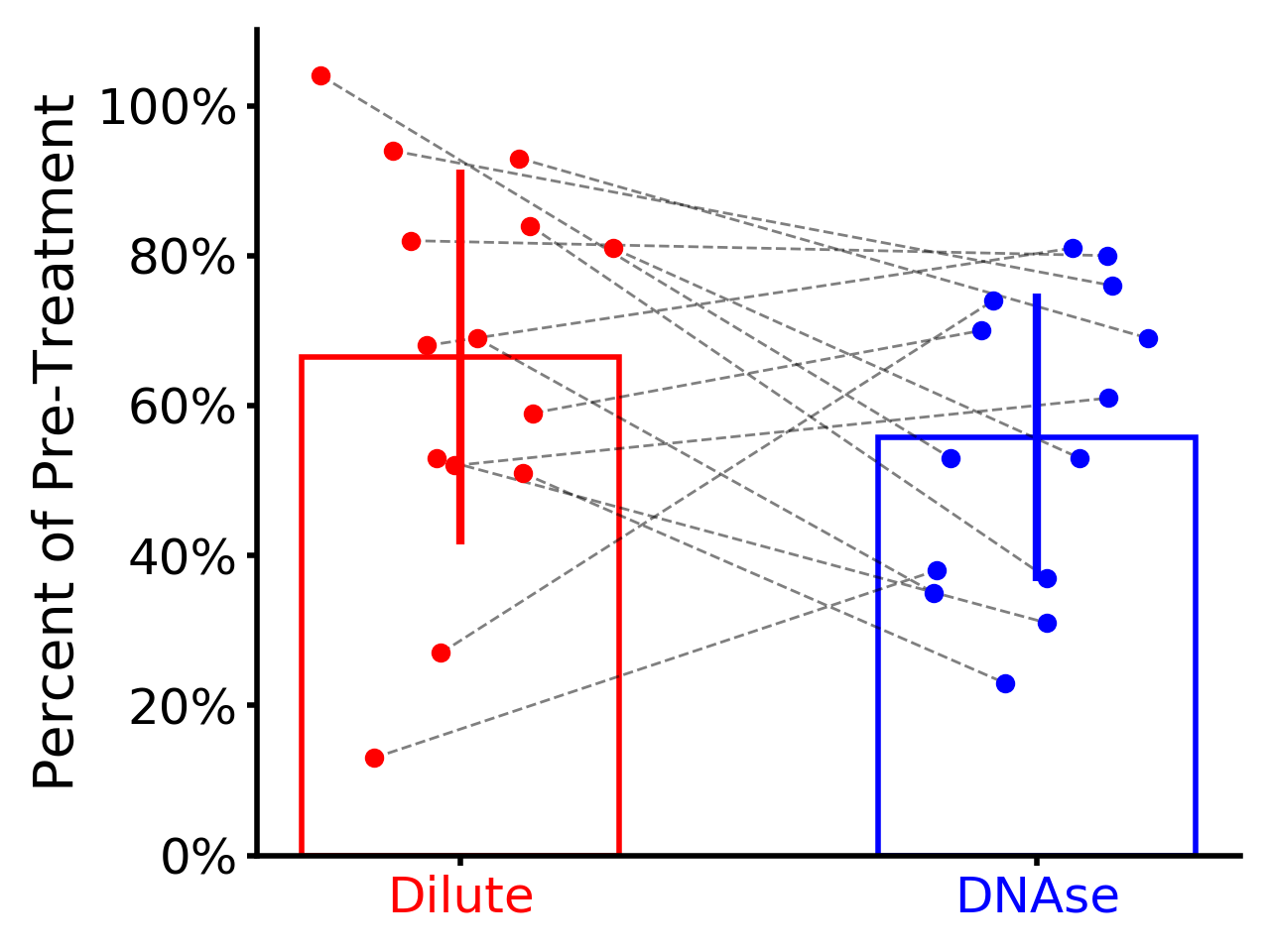

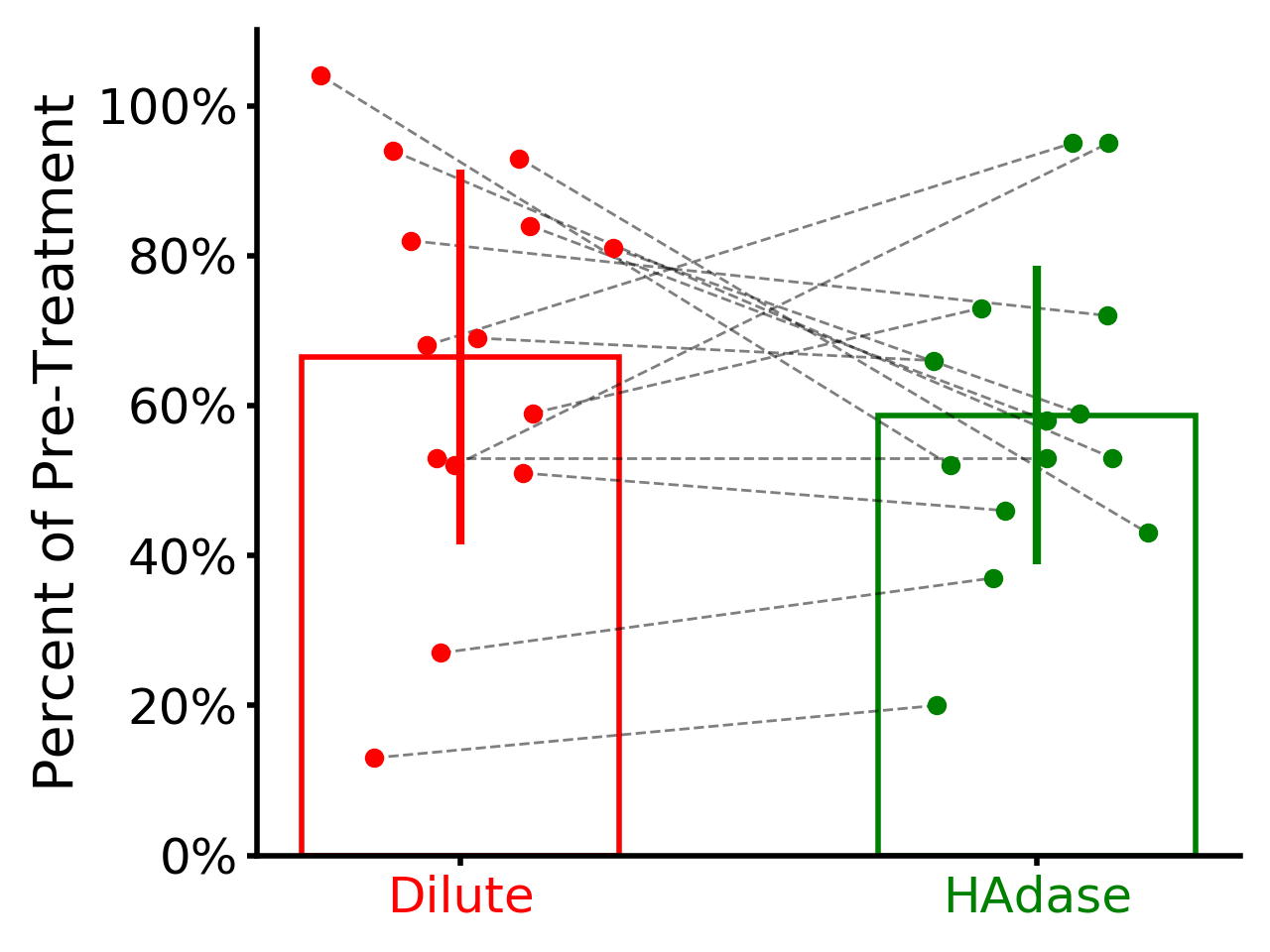

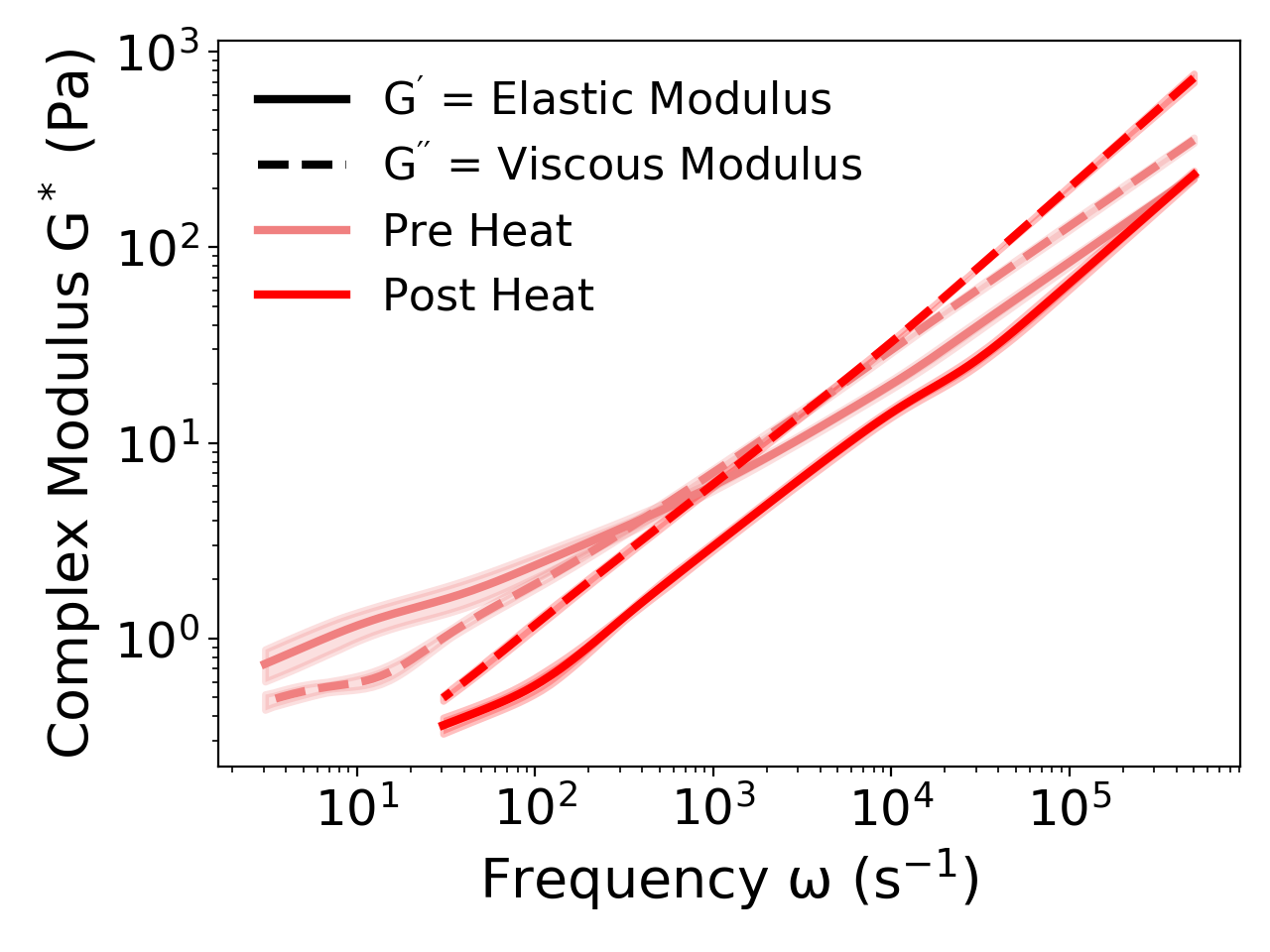

**A**

**B**

**C**

**D**

**Supplemental Figure 4.** (A). Microrheology data showing the impact of virus-killing heat treatment on cystic fibrosis (CF) respiratory secretions. (B) A plot of the measured moduli of all samples tested. Lighter and darker data points indicate the pre- and post-treatment, respectively, with a dashed line connecting data from the same patient sample. (C) A plot of relative change in modulus of COVID-19 respiratory secretions after dilution or HAdase enzymatic treatment compared to pre-treatment modulus, with a dashed line connecting data points generated from the same patient sample. (D) A plot of relative change in modulus of COVID-19 respiratory secretions after dilution or DNase enzymatic treatment compared to pre-treatment modulus, with a dashed line connecting data points generated from the same patient sample.

**Supplemental Acknowledgements**

The Stanford COVID-19 Biobank Study Group is: Rosen Mann, Anita Visweswaran, Thanmayi Ranganath, Jonasel Roque, Monali Manohar, Hena Naz Din, Komal Kumar, Kathryn Jee, Brigit Noon, Jill Anderson, Bethany Fay, Donald Schreiber, Nancy Zhao, Rosemary Vergara, Catherine Blish, Kari Nadeau, Andra Blomkalns, Ruth O'Hara.
